# Impact of intestinal parasitic infections on gut epithelial barrier and inflammation among foreign-born persons living with HIV

**DOI:** 10.1101/2025.02.03.25321616

**Authors:** Melissa Reimer-McAtee, Jose Serpa, Casey McAtee, Emma Ortega, Anoma Somasunderam, Roberto Arduino, Rojelio Mejia, Netanya S. Utay

## Abstract

Systemic inflammation is a major driver of comorbidities in people with HIV (PWH). Increased levels of biomarkers of enterocyte turnover, microbial translocation, and systemic inflammation have been shown to predict morbidity and mortality in PWH on antiretroviral therapy (ART). We conducted a prospective cohort study of foreign-born PWH with undetectable HIV RNA (<20 copies/mL) with and without intestinal parasitic coinfection. Biomarkers of enterocyte turnover (intestinal fatty acid binding protein [I-FABP]), microbial translocation (soluble CD14), and systemic inflammation (soluble CD163) were measured. Stool parasite quantitative PCR (qPCR) testing and *Strongyloides stercoralis* recombinant IgG ELISA (*Strongy* IgG) were utilized to diagnose parasitic infection. Of the 52 participants, 14 (27%) tested positive for infection with *Strongyloides stercoralis* by *Strongy* IgG, and 7 (16%) of the 45 participants who provided stool samples tested positive for a parasitic infection (not including *Blastocystis*) by stool qPCR. The median sCD14 level in PWH with (+) *Strongy* IgG was significantly higher than PWH with (-) *Strongy* IgG (1.69 ug/ml versus 1.48 ug/ml, p=0.03). The median sCD163 in PWH with parasitic infections by qPCR was not significantly different from sCD163 in PWH negative for parasitic infections. I-FABP levels did not differ significantly between groups. Participants with both HIV and intestinal parasite infections had increased levels of sCD14, a marker of microbial translocation that is an independent predictor of mortality in PWH, compared to PWH without parasitic infections. These findings raise concern about the long-term sequelae of intestinal parasitic infections in PWH.

## Introduction

Intestinal parasitic infections are often asymptomatic yet can cause chronic activation of immune responses in the human host^1–3^. Some parasites, such as *Strongyloides stercoralis*, can continuously autoinfect the host for the entire lifetime if left untreated^4,5^. Although treatable, intestinal parasitic infections are underdiagnosed due to lack of testing and low sensitivity of stool microscopy^6,7^. It is known that both HIV and enteric parasitic infection cause gut damage and increased microbial translocation^8–10^. Still, there are gaps in knowledge regarding the effects of simultaneous HIV and enteric parasitic infections on enterocyte turnover, microbial translocation, and systemic inflammation.

Parasitic infections increase CCR5^+^ CD4^+^ T-cell frequency, creating more HIV target cells in persons living with HIV (PWH)^10,11^. Parasitic infections also increase CD8^+^ T-cell activation^1^. Malnutrition, weight loss, chronic diarrhea, and changed immune responses caused by intestinal parasitic infections may further accelerate HIV/AIDS progression^12–16^. Reciprocally, HIV infection may impair the immune system’s ability to defend against parasitic infections^17,18^. Soluble CD14 (sCD14) is a marker of microbial translocation released upon LPS stimulation^19^. Both sCD14 and soluble CD163 (sCD163) are markers of macrophage and monocyte activation^19–21^. Intestinal fatty acid binding protein (I-FABP) is a marker of intestinal permeability and enterocyte turnover^20^. Increased levels of each of these biomarkers have been shown to predict morbidity and mortality in treated HIV infection, including cardiovascular disease and neurocognitive decline^19–25^.

We conducted a single-center, prospective cohort study to evaluate the impact of intestinal parasitic infections on microbial translocation (sCD14), systemic inflammation (sCD163), and enterocyte damage (I-FABP), and consequently, the potential for impacting morbidity and mortality of PWH.

## Materials and Methods

Participants were enrolled during a routine clinic visit at Thomas Street Health Center (TSHC), Harris Health System, the outpatient HIV center affiliated with Baylor College of Medicine and University of Texas Health Sciences Center-Houston. Inclusion criteria were as follows: age ≥18 years; foreign-born; documented HIV-1 infection; on antiretroviral therapy (ART) for >1 year; plasma HIV RNA < 20 copies/ml for ≥6 months (1 blip of < 200 copies/ml was permitted); and willingness to be treated with antiparasitic medication if indicated. Exclusion criteria were: inflammatory bowel disease; malignancy of the gastrointestinal tract; major intestinal surgery in the last 2 years; antiparasitic therapy in the last 1 year; pregnancy, plans to become pregnant, or breastfeeding; use of antibiotics in the last 60 days; use of probiotics and prebiotics (supplements and products) for more than three consecutive days within the 60 days prior to screening (yogurt with live cultures allowed). Ethical approval was obtained from Baylor College of Medicine and the University of Texas Health Sciences Center Institutional Review Boards. Administrative approval was granted by the Harris Health System Office of Research (H-37138). Written informed consent was obtained before any study procedures began.

Approximately 30g of stool and 20 mL of blood were collected from participants. A recombinant NIE-antigen ELISA was utilized to detect *Strongyloides stercoralis* infection, including positive and negative controls and a standard curve to quantify anti-*Strongyloides*-specific IgG^7^.

Multi-parallel real-time quantitative PCR (qPCR) was performed on stool to diagnose infection caused by *Ancylostoma duodenale, Ascaris lumbricoides, Blastocystis* species*, Cryptosporidium hominis/parvum, Cystoisospora belli, Entamoeba histolytica, Giardia intestinalis, Necator americanus, Trichuris trichiura, Schistosoma spp,* and *Strongyloides stercoralis*. Multi-parallel qPCR was performed as previously described^26^ with the same species-specific primers and FAM-labeled minor groove binder probes (Applied Biosystems, Foster City, CA). Samples were run on an ABI ViiA7 PCR in Houston, Texas, with default parameters and 40 cycles. DNA concentrations were calculated using a standard curve.

The biomarkers sCD14 and sCD163 were measured by Quantikine ELISA kit (R&D systems, MN), and I-FABP was measured by DuoSet ELISA kit (R&D systems, MN) according to the manufacturer’s instructions. The plasma samples were diluted as needed by the methods protocol.

We utilized stool quantitative PCR (qPCR) testing and *Strongyloides stercoralis* recombinant IgG ELISA (*Strongy* IgG) for sensitive diagnosis of parasitic infection to compare levels of sCD14, sCD163, and I-FABP to those without parasitic coinfection.

Results of these studies (qPCR and *Strongy* IgG) were reported to the participant’s clinician, who considered treatment according to the clinical condition of the patient and the local standard of care, as well as discussed risks and benefits with the participant. A medical chart review confirmed that each participant with positive *Strongy* IgG was treated with ivermectin. A follow-up evaluation of 6 months post-treatment was scheduled for each *Strongy* IgG positive participant, which consisted of a clinical evaluation and repeat blood samples for sCD14, sCD163, and I-FABP level testing.

Statistical analyses were performed with R^27^ and Prism 10.3.1 (GraphPad, Boston, MA). Correlations were estimated by calculating Spearman rank correlation. Mann-Whitney u-tests and Wilcoxon signed-rank tests were used to compare biomarker concentrations in parasite-infected and uninfected groups, pre-and post-treatment, respectively.

## Results

Fifty-two participants living with HIV born in 14 different countries were enrolled in February and March of 2019. Thirty-four of the 52 (65%) were male, and the median age of participants was 50 years (IQR 43-57). Participants had a median of 19 years living in the US **(Table 1)**. Median CD4^+^ T-cell count was 464 cells/mm^3^ (IQR 325-562). Plasma samples were obtained from all participants; stool samples were provided by 45 of the 52 participants. Fourteen (27%) participants tested positive for infection with *Strongyloides stercoralis* by *Strongy* IgG. Seven (15.5%) of the 45 participants who provided stool tested positive for pathogenic parasitic infection by stool qPCR: four (8.9%) with *Trichuris trichiura* (whipworm), one (2.2%) with *Necator americanus* (hookworm), one participant (2.2%) with *Strongyloides stercoralis,* and one (2.2%) with the protozoan infection *Entamoeba histolytica* **(Supplemental Figure 1)**. Three participants (7%) were coinfected with more than one parasitic infection (either *Strongy* IgG positive and stool PCR positive or stool PCR positive for two parasites). Of note, 37 (82.2%) tested positive for *Blastocystis* species, which for this study was considered non-pathogenic. All participants were negative for *Ancylostoma duodenale, Ascaris lumbricoides, Cryptosporidium hominis/parvum, Giardia intestinalis,* and *Schistosoma spp.* Seventeen of 52 (32.7%%) participants were positive for either *Strongy* IgG or stool qPCR (not including *Blastocystis* infection). One participant was positive for *Strongy* IgG and 3 parasites by stool qPCR-*Necator americanus* (hookworm), *Strongyloides stercoralis,* and *Trichuris trichiura* (whipworm). CD4^+^ T-cell counts were not significantly different between participants who tested positive for intestinal parasitic infection and those negative for parasitic infection (431 cells/mm^3^ [IQR 305-525] versus 492 cells/mm^3^ [IQR 320-655], respectively; p = 0.50). The median sCD14 level in (+) *Strongy* IgG participants was significantly higher than in (-) *Strongy* IgG participants (1.69 μg/ml [IQR 1.51-1.88)] versus 1.48 μg/ml [IQR 1.31-1.67], p = 0.03). The median sCD14 level was not significantly different between participants with or without parasitic infection (by *Strongy* IgG or stool PCR), not including *Blastocystis*: 1.69 μg/ml (IQR 1.23-2.51) compared to 1.38 μg/ml (0.78-4.22; p = 0.07) **(Figure 1)**. The median sCD163 level did not differ between (+) *Strongy* IgG vs (-) *Strongy* IgG participants (674 ng/ml [IQR 551-743] versus 583 ng/ml [IQR 480-760], p = 0.43). Similarly, the median sCD163 level did not differ between participants with parasitic infection (not including *Blastocystis*) versus parasite-negative participants (663 ng/ml [IQR 573-778] versus 569 ng/ml [471-721]; p = 0.20). I-FABP levels were not significantly different between (+) *Strongy* IgG and (-) *Strongy* IgG participants (832 pg/ml [IQR 380-1513] versus 921 pg/ml [IQR 362-2283], p = 0.98) or parasite-positive and parasite-negative participants (825 pg/ml [IQR 374-1898] versus 898 pg/ml [IQR 342-2214], p = 0.76). In a sensitivity analysis, participants positive and negative for parasites, including *Blastocystis* species, did not have statistically significant differences in any of the three biomarker levels.

**Figure 1.**
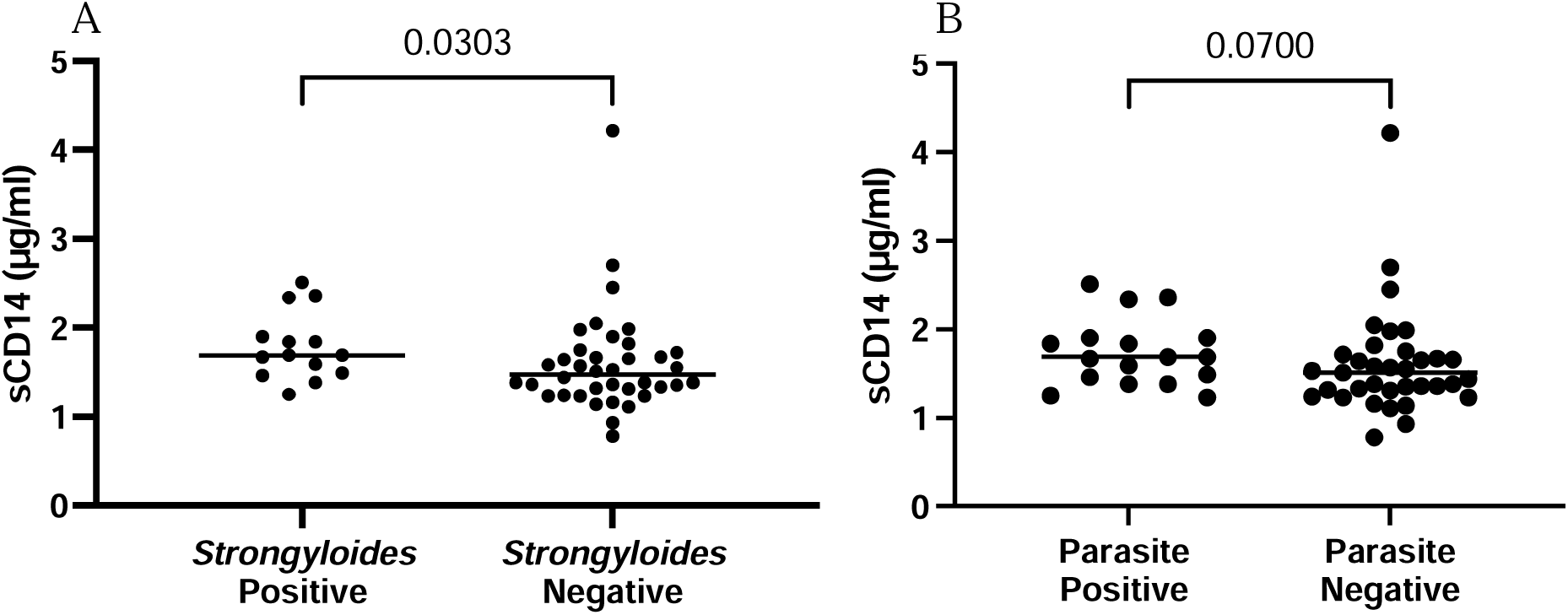
Soluble CD14 levels in those with and without parasites. Participants with strongyloidiasis had higher sCD14 levels (µg/ml) compared to participants without strongyloidiasis (1.69 vs. 1.48 µg/ml, respectively) (A). There was a higher sCD14 level (µg/ml) in individuals with either positive *Strongyloides* serology or positive qPCR for parasites not including *Blastocystis* (1.69 vs. 1.51 µg/ml, respectively) although not statistically significant (B). \**Strongyloides* positive indicates positive by *Strongyloides* IgG serology. **Parasite-positive indicates positive by either *Strongyloides* IgG serology or positive by stool PCR for pathogenic parasites

**Table 1.**
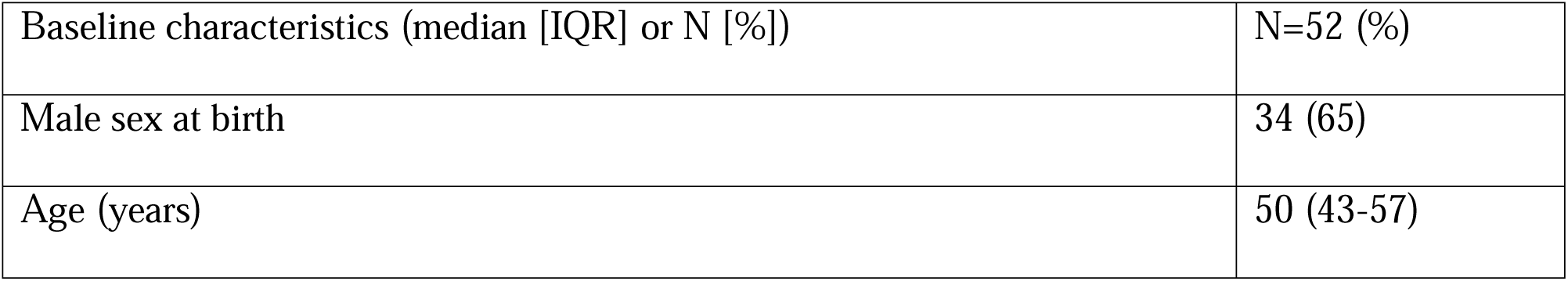
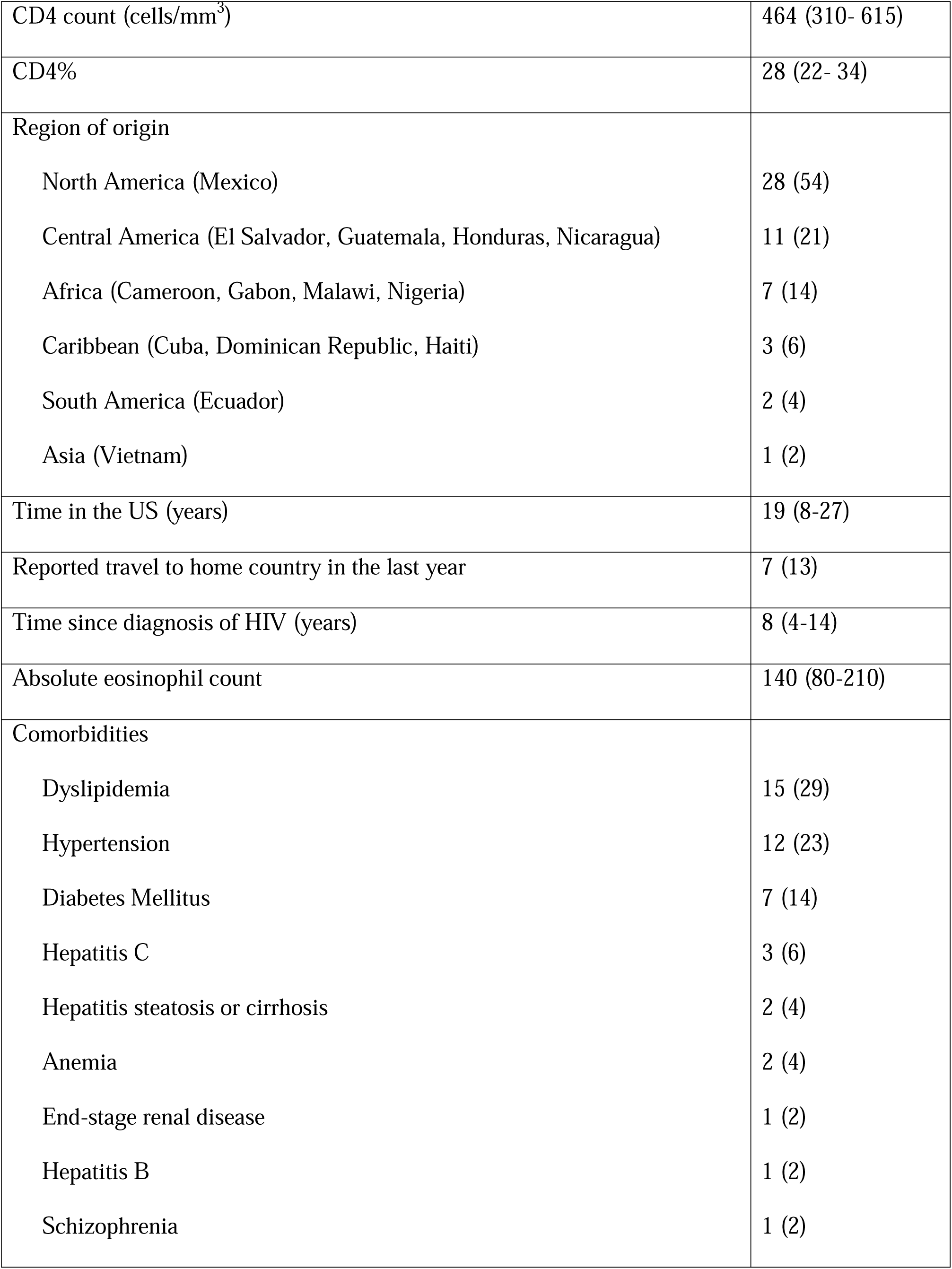
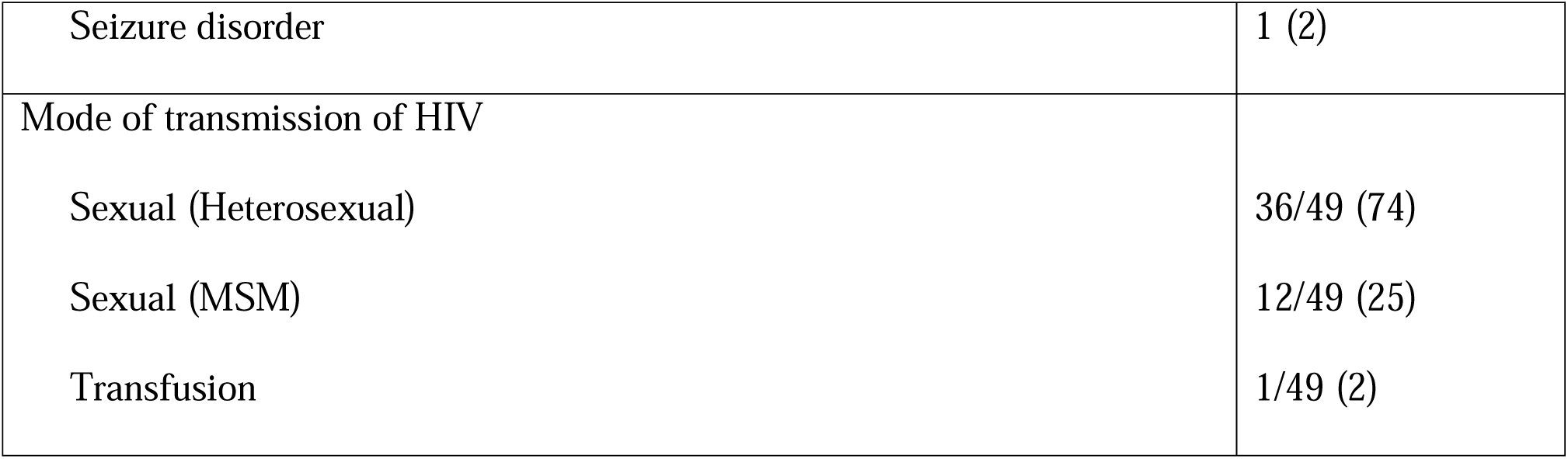
Patient Demographics.

| Baseline characteristics (median [IQR] or N [%]) | N=52 (%) |
| --- | --- |
| Male sex at birth | 34 (65) |
| Age (years) | 50 (43-57) |
| CD4 count (cells/mm<sup>3</sup>) | 464 (310- 615) |
| CD4% | 28 (22- 34) |
| Region of origin |  |
| North America (Mexico) | 28 (54) |
| Central America (El Salvador, Guatemala, Honduras, Nicaragua) | 11 (21) |
| Africa (Cameroon, Gabon, Malawi, Nigeria) | 7 (14) |
| Caribbean (Cuba, Dominican Republic, Haiti) | 3 (6) |
| South America (Ecuador) | 2 (4) |
| Asia (Vietnam) | 1 (2) |
| Time in the US (years) | 19 (8-27) |
| Reported travel to home country in the last year | 7 (13) |
| Time since diagnosis of HIV (years) | 8 (4-14) |
| Absolute eosinophil count | 140 (80-210) |
| Comorbidities |  |
| Dyslipidemia | 15 (29) |
| Hypertension | 12 (23) |
| Diabetes Mellitus | 7 (14) |
| Hepatitis C | 3 (6) |
| Hepatitis steatosis or cirrhosis | 2 (4) |
| Anemia | 2 (4) |
| End-stage renal disease | 1 (2) |
| Hepatitis B | 1 (2) |
| Schizophrenia | 1 (2) |
| Seizure disorder | 1 (2) |
| Mode of transmission of HIV |  |
| Sexual (Heterosexual) | 36/49 (74) |
| Sexual (MSM) | 12/49 (25) |
| Transfusion | 1/49 (2) |

Ten of the 14 participants (71%) positive by *Strongy* IgG on the initial visit were evaluated 6 months after parasitic infection treatment with post-treatment biomarker levels. Four of the 14 (29%) were lost to follow-up or unable to return because of restrictions due to the COVID-19 pandemic. Pre-treatment and post-treatment levels of sCD14 (1.69 vs. 1.735 µg/ml, p = 0.829), sCD163 (673 vs. 688 ng/ml, p = 0.752), and I-FABP (832.1 vs. 1418 pg/ml, p = 0.37) were not significantly different.

Logistical regression was used to detect predictors of *Strongy* IgG positivity. The number of years living in the United States, country of origin, years since visit to home country, and baseline absolute eosinophil count were not associated with test positivity.

## Discussion

Both HIV and parasitic infections are known to cause intestinal damage and affect inflammatory biomarkers, but the effects of HIV and parasite co-infection remain understudied. Our study provides a meaningful contribution to the literature because we evaluated levels of sCD14, sCD163, and I-FABP in PWH on suppressive ART in the setting of comprehensive testing for parasitic infections. We found that the median sCD14 level in PWH with (+) *Strongy* IgG was significantly higher than in PWH with (-) *Strongy* IgG. Given that sCD14 is an independent predictor of mortality in PWH^19^, the increased levels of sCD14 seen in participants of our study with parasitic infection raise concern about the long-term effects of enteric parasitic infection. The median sCD163 and I-FABP levels did not differ significantly between PWH with and without parasitic infections.

Our study detected a prevalence of 38% for pathogenic parasitic infection, demonstrating that intestinal parasitic coinfection is common in foreign-born PWH in the US. Most of these (27%) were due to infection with *Strongyloides stercoralis* detected by recombinant *Strongyloides* IgG ELISA. *Strongyloides stercoralis* infection is an auto-infectious helminth^4,5^. Thus, participants in our study could have been infected years before participation in our study. The *Strongyloides* adult worm releases only a few ova per day, making diagnosis difficult by stool microscopy and qPCR. A positive *Strongy* IgG can be considered a test for active infection in an untreated host^7^. Prior studies have shown that parasitic coinfection can negatively impact CD4 counts^28–31^ and plasma HIV RNA levels^32,33^, but most studies have evaluated PWH, not on suppressive ART. PWH in Southern Ethiopia not taking ART who were treated for *Ascaris lumbricoides* had a significant increase in CD4^+^ T-cells from 469 to 551 cells/mm^3^ at 6 months, compared to a non-significant decrease in helminth-uninfected participants^34^. Similar results were observed in a randomized, double-blind, placebo-controlled trial in ART-naive Kenyans treated for *Ascaris lumbricoides*^35^. PWH coinfected with hookworm diagnosed by stool qPCR in Uganda had significantly lower CD4^+^ T-cell counts, suggesting that intestinal parasitic infections could contribute to the low CD4^+^ T-cell count and delayed recovery in resource-limited settings^28^. Higher plasma HIV RNA levels were observed in pregnant women with hookworm or *Trichuris trichiura* infections^36^. Deworming of *T. trichiura* or hookworm in an observational study increased CD4^+^ T-cell counts and decreased the percentage of pregnant women on ART with detectable HIV RNA levels^37^.

In contrast to the studies above, a meta-analysis of randomized controlled trials of antiparasitic medications or placebo in PWH with unknown parasitic infection status showed no significant effect of one-time or repeated deworming on plasma HIV RNA levels or CD4^+^ T-cell count in people not taking ART^38^. A retrospective, observational study reported empiric deworming did not increase CD4^+^ T-cell counts among Ugandans on ART except when restricted to women in the first year of ART^39^. A study conducted in outpatient clinics in Lilongwe, Malawi, found no improvement in plasma HIV RNA levels after treatment of parasitic infection^40^.

It is important to note that most of the studies above used stool microscopy to identify those with parasitic infections. Given the low sensitivity of stool microscopy, the cohort of participants identified as parasite-negative may have included several participants with unidentified parasitic infections. Additionally, studies of parasitic infections in PWH *on suppressive ART* are limited. With the studies above reporting the variable impact of parasitic infection on markers of HIV infection, our study measured three biomarkers as an indirect evaluation of parasitic infection on HIV infection in coinfected participants. Soluble CD14 is a marker of microbial translocation and LPS-bioactivity^19^; both sCD14 and sCD163 are markers of monocyte and macrophage activation^19–21^. A case-control analysis of 74 PWH who died and 148 matched controls showed that participants with the highest quartile of soluble CD14 (sCD14) levels had a 6-fold increased risk of death compared to participants with the lowest quartile, even after adjusting for CD4^+^ T-cell count and other inflammatory markers^19^. The mechanism of this increase in mortality is not clear, but additional studies have shown an increase in cardiovascular disease and neurocognitive decline in patients with elevated biomarkers^20–23^, supporting the hypothesis that prolonged inflammation is harmful to the host, even if asymptomatic.

A key limitation of our study is the small sample size. Additionally, participants in our study left their home country at variable numbers of years before enrollment in the study, had various parasitic infections, and had blood, but no stool samples were available from some participants, which could have contributed to sampling error. Levels of sCD163 were higher in (+) *Strongy* IgG participants compared to (-) *Strongy* IgG participants and higher in participants positive by stool qPCR compared to participants negative by stool qPCR, but neither reached statistical significance. A study with a larger sample size would probably be needed further to study sCD163 levels in PWH with intestinal parasitic infections. We did not see a decrease in any of the three biomarkers measured following treatment of intestinal parasitic infection. This finding may have been related to our small sample size at follow-up visits, short-term follow-up, or true lack of improvement of LPS-bioactivity or change in monocyte and macrophage activation following treatment. Further studies are needed to determine the impact of treatment on these biomarkers. Another limitation is that we only had post-treatment biomarker results from the positive participants for *Strongy* IgG.

It is important to note that our study benefited from highly sensitive diagnostic tests that are not readily available outside the research setting. In most resource-limited clinical settings, the diagnosis of intestinal parasitic infection relies on poorly sensitive stool microscopy or empiric treatment is given based on clinical signs and symptoms. If additional studies show similar results with elevated levels of sCD14 and sCD163, or if a study with a larger post-treatment cohort shows improvement of biomarkers following treatment, then the clinical question of routine empiric deworming in PWH in parasite-endemic regions should be reconsidered. Given that sCD14 is an independent predictor of mortality in PWH, our study’s increased levels of sCD14 seen in PWH with parasitic infection raise concern about the long-term effects of enteric parasitic infection. Further studies are needed to determine the clinical and virological consequences of intestinal parasitic infections in PWH.

## Data Availability

All data produced in the present study are available upon reasonable request to the authors

## Acknowledgments

The authors wish to thank the the particpants and healthcare staff of this study.

## Disclosures regarding real or perceived conflicts of interest

None

## Co-Author Contact Information

1. Melissa Reimer-McAtee Division of Infectious Diseases, Department of Internal Medicine, University of Texas Health Sciences Center, Houston, TX, USA
2. Jose Serpa Division of Infectious Diseases, Department of Internal Medicine, University of Texas at Tyler, Tyler, TX, USA
3. Casey McAtee Department of Pediatrics, Baylor College of Medicine, Houston, TX, USA
4. Emma Ortega Department of Tropical Medicine and Infectious Disease, Tulane University School of Public Health and Tropical Medicine, New Orleans, LA, USA
5. Anoma Somasunderam Division of Infectious Diseases, Department of Internal Medicine, University of Texas Health Sciences Center, Houston, TX, USA
6. Roberto Arduino Division of Infectious Diseases, Department of Internal Medicine, University of Texas Health Sciences Center, Houston, TX, USA
7. Rojelio Mejia Department of Pediatrics: Tropical Medicine, Baylor College of Medicine, Houston, TX, USA
8. Netanya S. Utay Division of Infectious Diseases and Geographic Medicine, Department of Internal Medicine, University of Texas Southwestern Medical Center, Dallas, TX, USA

## References

1. Borkow G, Leng Q, Weisman Z, Stein M, Galai N, Kalinkovich A, Bentwich Z. Chronic immune activation associated with intestinal helminth infections results in impaired signal transduction and anergy. The Journal of clinical investigation. 2000;106(8):1053–60. Epub 2000/10/18. doi: 10.1172/jci10182. PubMed PMID: 11032865; PMCID: Pmc314342.

2. Ishikawa, N., P. K. Goyal, Y. R. Mahida, K. F. Li, and D. Wakelin. 1998. Early cytokine responses during intestinal parasitic infections. Immunology 93:257–263.

3. Loukas, A., and P. Prociv. 2001. Immune responses in hookworm infections. Clin. Microbiol. Rev. 14:689–703.

4. Page W, Judd JA, Bradbury RS. The Unique Life Cycle of Strongyloides stercoralis and Implications for Public Health Action. Trop Med Infect Dis. 2018 May 25;3(2):53. doi: 10.3390/tropicalmed3020053. PMID: 30274449; PMCID: PMC6073624.

5. Sing A, Leitritz L, Bogner JR, Heesemann J. First-glance diagnosis of Strongyloides stercoralis autoinfection by stool microscopy. J Clin Microbiol. 1999 May;37(5):1610–1. doi: 10.1128/JCM.37.5.1610-1611.1999. PMID: 10203537; PMCID: PMC84850.

6. Requena-Mendez A, Chiodini P, Bisoffi Z, Buonfrate D, Gotuzzo E, et al. (2013) The laboratory diagnosis and follow up of strongyloidiasis: a systematic review. PLoS Negl Trop Dis 7: e2002 10.1371/journal.pntd.0002002

7. Buonfrate D, Sequi M, Mejia R, Cimino RO, Krolewiecki AJ, Albonico M, Degani M, Tais S, Angheben A, Requena-Mendez A, Muñoz J, Nutman TB, Bisoffi Z. Accuracy of five serologic tests for the follow up of Strongyloides stercoralis infection. PLoS Negl Trop Dis. 2015 Feb 10;9(2):e0003491. doi: 10.1371/journal.pntd.0003491. PMID: 25668740; PMCID: PMC4323101.

8. Brenchley JM, Price DA, Schacker TW, Asher TE, Silvestri G, Rao S, et al.. Microbial translocation is a cause of systemic immune activation in chronic HIV infection. Nat Med (2006) 12(12):1365–71. doi: 10.1038/nm1511

9. McKay DM, Shute A, Lopes F. Helminths and intestinal barrier function. Tissue Barriers. 2017 Jan 2;5(1):e1283385. doi: 10.1080/21688370.2017.1283385. PMID: 28452686; PMCID: PMC5362995.

10. George PJ, Anuradha R, Kumar NP, Kumaraswami V, Nutman TB, Babu S. Evidence of microbial translocation associated with perturbations in T cell and antigen-presenting cell homeostasis in hookworm infections. PLoS neglected tropical diseases. 2012;6(10):e1830. Epub 2012/10/12. doi: 10.1371/journal.pntd.0001830. PubMed PMID: 23056659; PMCID: Pmc3464301.

11. Sher A, Fiorentino D, Caspar P, Pearce E, Mosmann T. Production of IL-10 by CD4+ T lymphocytes correlates with down-regulation of Th1 cytokine synthesis in helminth infection. Journal of immunology (Baltimore, Md : 1950). 1991;147(8):2713–6. Epub 1991/10/15. PubMed PMID: 1680917.

12. Sangare LR, Herrin BR, John-Stewart G, Walson JL. Species-specific treatment effects of helminth/HIV-1 co-infection: a systematic review and meta-analysis. Parasitology. 2011;138(12):1546–58. Epub 2011/07/07. doi: 10.1017/s0031182011000357. PubMed PMID: 21729353;

13. Onguru D, Liang Y, Griffith Q, Nikolajczyk B, Mwinzi P, Ganley-Leal L. Human schistosomiasis is associated with endotoxemia and Toll-like receptor 2- and 4-bearing B cells. The American journal of tropical medicine and hygiene. 2011;84(2):321–4. Epub 2011/02/05. doi: 10.4269/ajtmh.2011.10-0397. PubMed PMID: 21292908; PMCID: Pmc3029191.

14. Aksoy E, Zouain CS, Vanhoutte F, Fontaine J, Pavelka N, Thieblemont N, Willems F, Ricciardi-Castagnoli P, Goldman M, Capron M, Ryffel B, Trottein F. Double-stranded RNAs from the helminth parasite Schistosoma activate TLR3 in dendritic cells. The Journal of biological chemistry. 2005;280(1):277–83. Epub 2004/11/03. doi: 10.1074/jbc.M411223200. PubMed PMID: 15519998.

15. Turner JD, Jenkins GR, Hogg KG, Aynsley SA, Paveley RA, Cook PC, Coles MC, Mountford AP. CD4+CD25+ regulatory cells contribute to the regulation of colonic Th2 granulomatous pathology caused by schistosome infection. PLoS neglected tropical diseases. 2011;5(8):e1269. Epub 2011/08/23. doi: 10.1371/journal.pntd.0001269. PubMed PMID: 21858239; PMCID: Pmc3153428.

16. Singh KP, Gerard HC, Hudson AP, Boros DL. Differential expression of collagen, MMP, TIMP and fibrogenic-cytokine genes in the granulomatous colon of Schistosoma mansoni-infected mice. Annals of tropical medicine and parasitology. 2006;100(7):611–20 Epub 2006/09/23. doi: 10.1179/136485906x118530. PubMed PMID: 16989687.

17. Ramakrishnan K, Shenbagarathai R, Uma A, Kavitha K, Rajendran R, Thirumalaikolundusubramanian P. Prevalence of intestinal parasitic infestation in HIV/AIDS patients with diarrhea in Madurai City, South India. Japanese journal of infectious diseases. 2007;60(4):209–10. Epub 2007/07/24. PubMed PMID: 17642535.

18. Tian LG, Wang TP, Lv S, Wang FF, Guo J, Yin XM, Cai YC, Dickey MK, Steinmann P, Chen JX. HIV and intestinal parasite co-infections among a Chinese population: an immunological profile. Infectious diseases of poverty. 2013;2(1):18. Epub 2013/08/27. doi: 10.1186/2049-9957-2-18. PubMed PMID: 23971713; PMCID: Pmc3766051.

19. Sandler NG, Wand H, Roque A, Law M, Nason MC, Nixon DE, Pedersen C, Ruxrungtham K, Lewin SR, Emery S, Neaton JD, Brenchley JM, Deeks SG, Sereti I, Douek DC. Plasma Levels of Soluble CD14 Independently Predict Mortality in HIV Infection. J Infect Dis. 2011. Epub 2011/01/22. doi: jiq118 [pii]10.1093/infdis/jiq118. PubMed PMID: 21252259.

20. Hunt PW, Sinclair E, Rodriguez B, Shive C, Clagett B, Funderburg N, Robinson J, Huang Y, Epling L, Martin JN, Deeks SG, Meinert CL, Van Natta ML, Jabs DA, Lederman MM. Gut epithelial barrier dysfunction and innate immune activation predict mortality in treated HIV infection. J Infect Dis. 2014;210(8):1228–38. doi: 10.1093/infdis/jiu238. PubMed PMID: 24755434; PMCID: PMC4192038.

21. Kelesidis T, Kendall MA, Yang OO, Hodis HN, Currier JS. Biomarkers of microbial translocation and macrophage activation: association with progression of subclinical atherosclerosis in HIV-1 infection. J Infect Dis. 2012;206(10):1558–1567. doi:10.1093/infdis/jis545

22. Tenorio AR, Zheng Y, Bosch RJ, et al. Soluble markers of inflammation and coagulation but not T-cell activation predict non-AIDS-defining morbid events during suppressive antiretroviral treatment. J Infect Dis. 2014;210(8):1248–1259. doi:10.1093/infdis/jiu254

23. Longenecker CT, Jiang Y, Orringer CE, et al. Soluble CD14 is independently associated with coronary calcification and extent of subclinical vascular disease in treated HIV infection. AIDS. 2014;28(7):969–977. doi:10.1097/QAD.0000000000000158

24. Knudsen T, Ertner G, Petersen J, et al. Plasma Soluble CD163 Level Independently Predicts All-Cause Mortality in HIV-1–Infected Individuals, The Journal of Infectious Diseases, Volume 214, Issue 8, 15 October 2016, Pages 1198–1204, 10.1093/infdis/jiw263

25. Utay NS, Overton ET. Immune Activation and Inflammation in People With Human Immunodeficiency Virus: Challenging Targets. J Infect Dis. 2020 Apr 27;221(10):1567–1570. doi: 10.1093/infdis/jiz351. PMID: 31282534; PMCID: PMC7184903.

26. Mejia R, Vicuña Y, Broncano N, et al. A novel, multi-parallel, real-time polymerase chain reaction approach for eight gastrointestinal parasites provides improved diagnostic capabilities to resource-limited at-risk populations. Am J Trop Med Hyg. 2013;88(6):1041–1047. doi:10.4269/ajtmh.12-0726

27. R Core Team (2021). R: A language and environment for statistical computing. R Foundation for Statistical Computing, Vienna, Austria. URL https://www.R-project.org/

28. Morawski BM, Yunus M, Kerukadho E, et al. Hookworm infection is associated with decreased CD4+ T cell counts in HIV-infected adult Ugandans. PLoS Negl Trop Dis. 2017;11(5):e0005634. Published 2017 May 25. doi:10.1371/journal.pntd.0005634

29. Kalinkovich A, Weisman Z, Greenberg Z, Nahmias J, Eitan S, Stein M, Bentwich Z. Decreased CD4 and increased CD8 counts with T cell activation is associated with chronic helminth infection. Clinical and experimental immunology. 1998;114(3):414–21. Epub 1998/12/09. PubMed PMID: 9844052; PMCID: Pmc1905129.

30. Arndt MB, John-Stewart G, Richardson BA, Singa B, van Lieshout L, Verweij JJ, Sangare LR, Mbogo LW, Naulikha JM, Walson JL. Impact of helminth diagnostic test performance on estimation of risk factors and outcomes in HIV-positive adults. PloS one. 2013;8(12):e81915. Epub 2013/12/11. doi: 10.1371/journal.pone.0081915. PubMed PMID: 24324729; PMCID: Pmc3852669.

31. Walson JL, Stewart BT, Sangare L, Mbogo LW, Otieno PA, Piper BK, Richardson BA, John-Stewart G. Prevalence and correlates of helminth co-infection in Kenyan HIV-1 infected adults. PLoS neglected tropical diseases. 2010;4(3):e644. Epub 2010/04/03. doi: 10.1371/journal.pntd.0000644. PubMed PMID: 20361031; PMCID: Pmc2846937.

32. Turner JD, Jenkins GR, Hogg KG, Aynsley SA, Paveley RA, Cook PC, Coles MC, Mountford AP. CD4+CD25+ regulatory cells contribute to the regulation of colonic Th2 granulomatous pathology caused by schistosome infection. PLoS neglected tropical diseases. 2011;5(8):e1269. Epub 2011/08/23. doi: 10.1371/journal.pntd.0001269. PubMed PMID: 21858239; PMCID: Pmc3153428.

33. Singh KP, Gerard HC, Hudson AP, Boros DL. Differential expression of collagen, MMP, TIMP and fibrogenic-cytokine genes in the granulomatous colon of Schistosoma mansoni-infected mice. Annals of tropical medicine and parasitology. 2006;100(7):611–20 Epub 2006/09/23. doi: 10.1179/136485906x118530. PubMed PMID: 16989687.

34. Abossie A, Petros B. Deworming and the immune status of HIV positive pre-antiretroviral therapy individuals in Arba Minch, Chencha and Gidole hospitals, Southern Ethiopia. BMC research notes. 2015;8:483. Epub 2015/09/30. doi: 10.1186/s13104-015-1461-9. PubMed PMID: 26415705; PMCID: Pmc4585997

35. Walson JL, Otieno PA, Mbuchi M, Richardson BA, Lohman-Payne B, Macharia SW, Overbaugh J, Berkley J, Sanders EJ, Chung MH, John-Stewart GC. Albendazole treatment of HIV-1 and helminth co-infection: a randomized, double-blind, placebo-controlled trial. AIDS (London, England). 2008;22(13):1601–9. Epub 2008/08/02. doi: 10.1097/QAD.0b013e32830a502e. PubMed PMID: 18670219; PMCID: Pmc2637615.

36. Webb EL, Kyosiimire-Lugemwa J, Kizito D, Nkurunziza P, Lule S, Muhangi L, Muwanga M, Kaleebu P, Elliott AM. The effect of anthelmintic treatment during pregnancy on HIV plasma viral load: results from a randomized, double-blind, placebo-controlled trial in Uganda. J Acquir Immune Defic Syndr. 2012 Jul 1;60(3):307–13. doi: 10.1097/QAI.0b013e3182511e42. PMID: 22728750; PMCID: PMC3383620.

37. Ivan E, Crowther NJ, Mutimura E, Rucogoza A, Janssen S, Njunwa KK, Grobusch MP. Effect of deworming on disease progression markers in HIV-1-infected pregnant women on antiretroviral therapy: a longitudinal observational study from Rwanda. Clinical infectious diseases : an official publication of the Infectious Diseases Society of America. 2015;60(1):135–42. Epub 2014/09/12. doi: 10.1093/cid/ciu715. PubMed PMID: 25210019.

38. Means AR, Burns P, Sinclair D, Walson JL. Antihelminthics in helminth-endemic areas: effects on HIV disease progression. Cochrane Database Syst Rev. 2016 Apr 14;4(4):CD006419. doi: 10.1002/14651858.CD006419.pub4. PMID: 27075622; PMCID: PMC4963621.

39. Lankowski AJ, Tsai AC, Kanyesigye M, Bwana M, Haberer JE, Wenger M, Martin JN, Bangsberg DR, Hunt PW, Siedner MJ. Empiric deworming and CD4 count recovery in HIV-infected Ugandans initiating antiretroviral therapy. PLoS neglected tropical diseases. 2014;8(8):e3036. Epub 2014/08/08. doi: 10.1371/journal.pntd.0003036. PubMed PMID: 25101890; PMCID: Pmc4125278.

40. Hosseinipour MC, Napravnik S, Joaki G, Gama S, Mbeye N, Banda B, Martinson F, Hoffman I, Cohen MS. HIV and parasitic infection and the effect of treatment among adult outpatients in Malawi. J Infect Dis. 2007 May 1;195(9):1278–82. doi: 10.1086/513274. Epub 2007 Mar 20. PMID: 17396996.

