## Supplemental Figure 1. for "Impact of intestinal parasitic infections on gut epithelial barrier and inflammation among foreign-born persons living with HIV"

Supplemental material

**Supplemental Figure 1. Prevalence and DNA burden of parasites (N, %).** 2.2% of participants were positive for *N. americanus, S. stercoralis,* and *E. histolytica* and a concentration of parasite DNA (3.09, 0.316, and 0.579 fg/µl, respectively). *Blastocystis* species had a mean of 696.6 fg/µl and 82% positivity. *T. trichiura* had a mean 0.0322 fg/µl and 8.9% positivity.

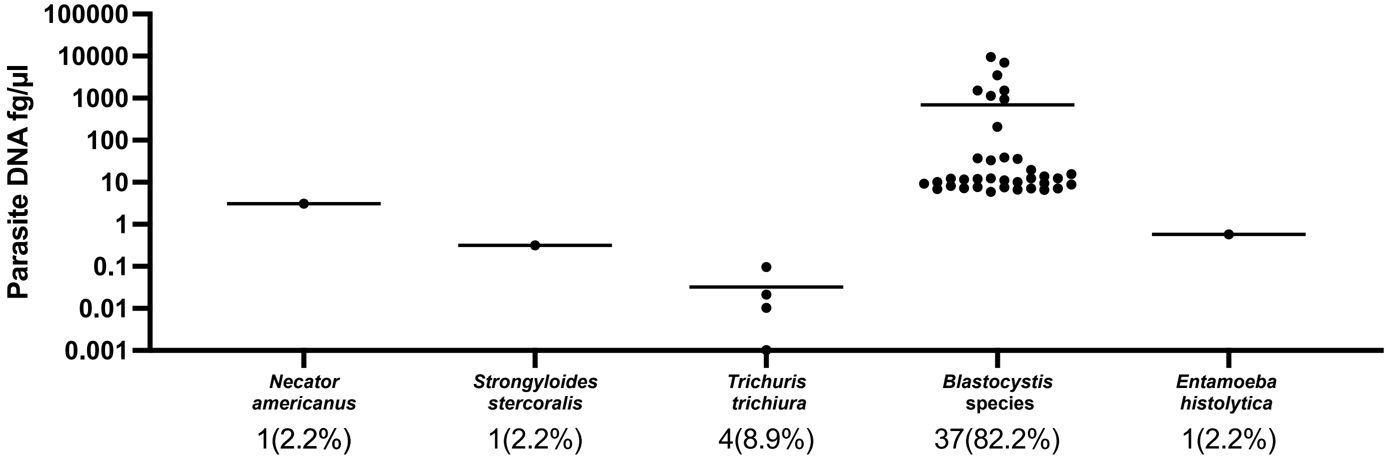
